# Use of artificial intelligence on retinal images to accurately predict the risk of cardiovascular event (CVD-AI)

**DOI:** 10.1101/2022.10.12.22281017

**Authors:** Ehsan Vaghefi, David Squirrell, Song Yang, Songyang An, John Marshall

**Affiliations:** Toku Eyes, Auckland, New Zealand; School of Ophthalmology, University College London, United Kingdom

## Abstract

**Purpose:** To create and evaluate the accuracy of an artificial intelligence platform capable of using only retinal fundus images to predict both an individual*’*s overall 10 year Cardiovascular Disease (CVD) risk and the relative contribution of the component risk factors that comprise this risk (CVD-AI).

**Methods:** The UK Biobank and the US-based AREDS 1 datasets were obtained and used for this study. The UK Biobank data was used for training, validation and testing, while the AREDS 1 dataset was used as an external testing dataset. Overall, we used 110,272 fundus images from 55,118 patient visits. A series of models were trained to predict the risk of CVD against available labels in the UK Biobank dataset.

**Results:** In both the UK Biobank testing dataset and the external validation dataset (AREDS 1), the 10-year CV risk scores generated by CVD-AI were significantly higher for patients who had suffered an actual CVD event when compared to patients who did not experience a CVD event. In the UK Biobank dataset the median 10-year CVD risk for those individuals who experienced a CVD was higher than those who did not (4.9% [ICR 2.9-8%] v 2.3% [IQR 4.3-1.3%] P<0.01.]. Similar results were observed in the AREDS 1 dataset The median 10-year CVD risk for those individuals who experienced a CVD event was higher than those who did not (6.2% [ICR 3.2%-12.9%] v 2.2% [IQR 3.9-1.3%] P<0.01

**Conclusion:** Retinal photography is inexpensive and as fully automated, inexpensive camera systems are now widely available, minimal training is required to acquire them. As such, AI enabled retinal image-based CVD risk algorithms like CVD-AI will make cardiovascular health screening more affordable and accessible. CVD-AI*’*s unique ability to assess the relative contribution of the components that comprise an individual*’*s overall risk could inform personalized treatment decisions based on the specific needs of an individual, thereby increasing the likelihood of positive health outcomes.

## Background

Cardiovascular disease (CVD) is the commonest cause of hospitalization and premature death in the US [1]. The risk of an individual experiencing a CVD event includes both non-modifiable; age, sex, and ethnicity, and modifiable variables such as diabetes [2], hypertension [3], hyperlipidemia [4], and smoking [5]. Across a population the risk of experiencing a CVD event varies greatly and risk based equations have therefore been developed to identify those who are greatest risk of a CVD event risk so that treatments can be instigated appropriate to the individuals risk [6].

The landmark Framingham Heart Study was the first to demonstrate that multivariable equations could identify an individual*’*s CVD risk with far greater accuracy than the existing metrics based solely on blood pressure and cholesterol [7]. Since the Framingham-based equations were first published other equations have been developed designed to serve different populations with refined accuracy [8-15]. Kavousi et al reviewed the differences in prevention risk scores between three different guidelines (American College of Cardiology/American Heart Association, the Adult Treatment Panel III, and the European Society of Cardiology guidelines) on a sample of 4854 participants from the Netherlands. They found that these three models provided poor calibration and only moderate to good discrimination between subjects and their findings highlight the importance of both continuing to improve risk predictions and setting appropriate population-wide thresholds [8].

Although imperfect, the use of multivariable equations has not only improved the accuracy of calculating an individual*’*s CVD risk, they have also improved our understanding of the complex interplay of factors that underpin this risk. This statistical approach does however have limitations. A recent systematic review and meta-analysis found that the Framingham-based risk models and pooled cohort equations for predicting 10-year risk of CVD not only had a tendency to overestimate the risk level, especially in higher-risk populations [16], they prove unreliable for people living with diabetes [17]. One fundamental weakness of all existing multivariant equations is that the predictors used are not a direct measure of CVD. Instead the equations are based on regression models which utilize parameters known to correlate with CVD, including age, sex, ethnicity, socioeconomic deprivation, smoking, diabetes duration, systolic blood pressure (SBD), total cholesterol-to-HDL ratio (TCHDL) and glycated hemoglobin A1c (HbA1c) [18]. This approach is limited by the fact that the strength of these correlations will differ between groups, and, as such, the predictive power of the equation will vary depending on the clinical profile of a local population and a given individual within it [19]. This probably explains why the generic CVD risk equations often do not perform as well as models developed for specific populations (10). Additionally, the majority of CVD risk equations developed to date attempt to identify patients who are at risk of experiencing cardiovascular events based on data obtained within a specific period of time. Invariably not all data is available at all time points [20] and the issue of missing data means that the equations may not accurately reflect risk in those who do not engage with, or have access to healthcare services [21].

The retina is unique being the only part of the human vasculature that is visible by non-invasive means. In recent years there has been an exponential increase in the number of studies that have used Artificial intelligence (AI), and deep learning (DL) in particular, to extract data from retinal images [22]. Having recognized the power of DL to extract data from retinal images there is now intense interest in using the retinal image data generated by DL algorithms to augment the traditional means of estimating CVD risk [23]. To date algorithms have been developed that generate a modifier to an individual chronological age to predict their biological Cardiovascular age [24], coronary artery calcium (CAC) scores [25], CT-based coronary artery disease [26], and those modifiable and non-modifiable CVD risk factors that contribute to CVD risk [27]. DL algorithms have also been developed recently that aim to predict CVD risk directly [28]. Others have taken a slightly different approach using genome-wide association studies to investigate the genetic component of retinal vasculature measured as fractal dimension analyzing its relationship with CVD [29]. More recently, Cheung et al described a DL model for the assessment of cardiovascular disease risk via the measurement of retinal-vessel caliber [25]. The model was trained on multiethnic multi-country datasets that comprised more than 70,000 retinal images and provided performance that was comparable to, or better than, expert graders in associations between measurements of retinal-vessel caliber and CVD risk factors (e.g., blood pressure, body-mass index, cholesterol and glycated-hemoglobin levels).

We have developed a DL model (CVD-AI), that predicts an individual*’*s 10-year CVD risk directly based solely on their retinal photograph. Unlike traditional CVD risk equations and existing deep learning prediction models, CVD-AI calculates the interactions between modifiable factors when assessing the risk contribution of each to the total risk score. By doing so CVD-AI learns if changes to one modifiable factor correlate with changes in other modifiable factors. In this study, we used 110,272 fundus images from a database of 55,118 patients to calculate a CVD risk score based on our platform CVD-AI. We then compared the results with the actual cardiovascular event rate to determine the efficacy of the prediction methods.

## Methods

### Data preparation

In total, 110,272 fundus images from a database of 55,118 patient visits were used in this study. The composition of the data can be accessed here https://public.tableau.com/app/profile/toku.eyes/viz/UK_Biobank/Story?publish=yes. & https://public.tableau.com/app/profile/toku.eyes/viz/AREDS/Story?publish=yes The dataset was acquired from the UK Biobank and AREDS 1, using approved data management and data transfer protocols.

From the UK Biobank dataset, initially 175,788 macula centered images for both the left and right eyes were acquired. However, due to the prevalence of low-quality images, a deep learning image quality screening system was used to separate the images into high quality, medium quality, and low-quality images. The training dataset for the image quality screening system was in a similar manner to our prior THEIA system [30]. After the screening process there were 95,992 images from 51,956 patients (Figure 1). Due to patients visiting Biobank for repeated assessment visits, there were multiple sets of biometric information per patient. Here, only the earliest images and set of biometrics were used per patient.

**Figure 1:**
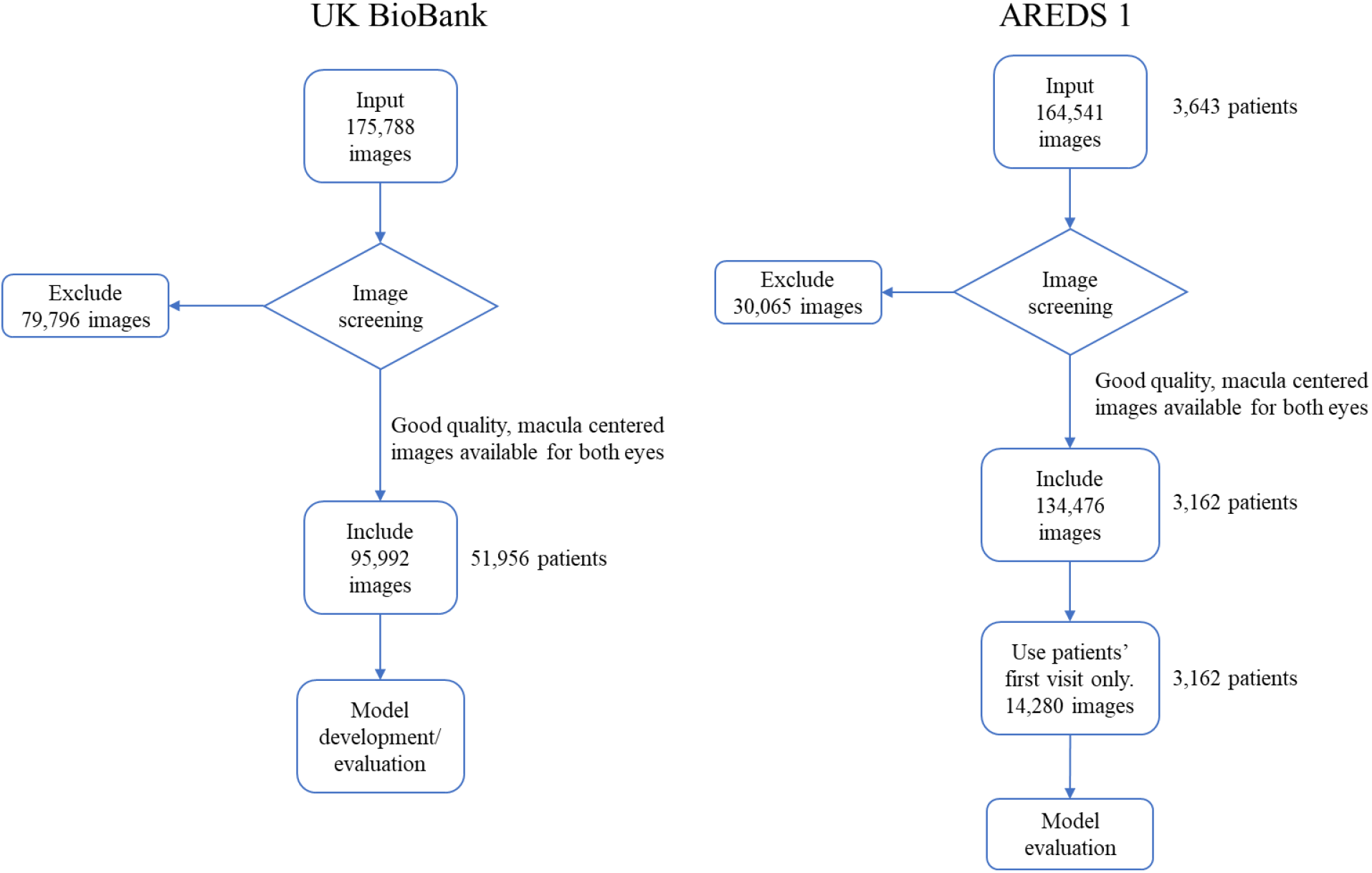
data preparation and refinement from UK Biobank and AREDS 1 datasets, to be included in this study

Dataset processing was also carried out on the AREDS 1 dataset. A similar image screening pre-processing strategy was employed for the AREDS 1 dataset, and the biometric information and fundus images from the first visit only. After screening for image quality, there were 134,476 images from 3,162 patients left for test analysis (Figure 1).

To generate a label for the presence or absence of CVD events, ICD10/ICD9 codes were utilized. For the UK Biobank dataset, CVD events were defined based on the *“*Diagnoses – ICD10*”* and *“*Underlying (primary) cause of death: ICD10*”* fields. For the AREDS dataset, CVD events were defined based on ICD 9 codes from the *“*ICD9COD1*”* variable under the *“*adverse*”* dataset and the *“*ICD10*”* and *“*ICD9COD1*”* variables under the mortality dataset. The criterion for the ICD10 representing CVD events was obtained upon request from the PREDICT study [31]. A detailed overview of the demographic distribution of the final processed UK Biobank and AREDS 1 datasets can be found in the supplementary materials [Supplementary Tables 1&2].

### Model Development

The UK Biobank dataset (95,992 good quality images from 51,956 patients) was split into 70%:15%:15% for training, validation and testing respectively. For the external test set, AREDS 1 dataset (14,280 good quality images from 3,162 patients) was used. To extract features from the fundus images for the Convolutional neural networks (CNN)s base, we created an ensemble of CNNs to look at a number of datapoints in the fundus image. These CNNs follow modified versions of the Inception-Resent-V2 or ResNet50 structures. For each CNN, the dataset was then split for training, validation, and testing. The fundus images were first cropped and resized to 800×800 pixel size. The batch size was set to 8 to optimize the use of the GPU memory during training. Adam optimizer was adopted with a learning rate 1*10e-3 to update parameters towards the minimization of the loss. Dropout was enabled with a rate p = 0.2, and the model was trained for at least 100 EPOCHs. All codes related to this work were implemented by Python 3.7. programming language.

The extracted image features from the ensemble of CNNs were fed into the final risk prediction model along with the patient*’*s age, gender, ethnicity. The cross-entropy loss function was employed to guide the model parameters optimization. The training objective was to minimize the loss function to get the most accurate probability prediction of CVD events. Typically, cross-entropy loss is utilized in classification problems. Although the CVD event risk prediction is not a classification task, the label used in this study was either 1 or 0, indicating whether a CVD event happened or not, respectively. By doing so we adopted the cross-entropy loss. The overall loss can be formalized as

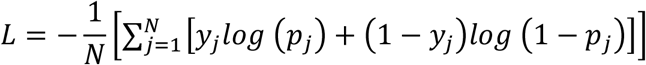

where *N* is the number of training samples, *yj* is the ground truth of sample j, and *pj* is the predicted probability of CVD for sample j. The model optimization is guided by minimizing the cross-entropy loss, which is a representation of the amount of divergence between the distributions of the labels and the predicted probabilities.

### Model explainability

To gain a better insight into the behavior of the final multilayer perceptron predictor, the expected gradients algorithm [32] was implemented to estimate the absolute contribution of each of the inputs to the CVD risk multilayer perceptron risk predictor. The expected gradients algorithm estimates an *“*attribution score*”* for each of the input fields such as age, gender, systolic blood pressure that are used by the multilayer perceptron risk predictor model to estimate the final CVD risk score. The *“*attribution score*”* for an input field, such as age, represents the amount of difference this particular field contributed to the value difference between the predicted CVD risk for this particular patient and that of the entire source dataset. The attribution score for the following key fields were calculated as these factors have been identified as the major contributing factors to an individual cardiovascular risk by the American College of Cardiology[33] :

1. Age
2. Gender
3. BMI
4. Smoking status (represented by model predicted smoking status)
5. Glycemic control (represented by model predicted *“*effect*”* of HbA1C)
6. Blood pressure (represented by model predicted *“*effect*”* of systolic and Diastolic blood pressure)
7. Cholesterol/ HDL (represented by model predicted *“*effect*”* of TCHDL ratio)

The attribution scores for all other input fields into the CVD risk prediction model were categorized and summated under the *“*others aggregated*”* category.

Due to the uneven magnitudes between the attribution scores for different prediction cases, for example, the attribution scores for a patient with 20% CVD-AI predicted risk will of far greater magnitude when compared to a patient with 12% CVD-AI predicted risk, a scaling algorithm shown in the equation below was used to scale the attribution scores between 0 and 100%.

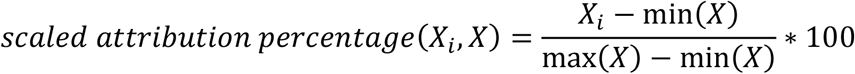

In this case, *X* represents the set of attribution scores, and *Xi* represents an element of the set of attribution scores.

Having calculated the attribution scores generated by the modified expected gradients the plausibility of the outputs produced by CVD-AI, based simply on the retinal image, were evaluated in three ways in both the internal validation dataset (Biobank) and the external validation dataset (AREDS 1).

### Primary outcome

Comparison of population based mean and median scores CVD-AI allocated to those individuals who actually experienced a CVD event, compared to those who did not.

### Secondary outcomes

1. By way of comparing the demographics and biometric data of those individuals that CVD-AI categorized as low, medium and high risk; defined by CV risk assessments scores of <5% 5-10% and over 10% respectively.[33-35]

2. By way of a set of case studies to qualitatively compare the attribution scores generated by CVD-AI to the real-life status of individual patients.

### Statistical analysis

The following statistical methods were applied to analyze the data. The specific method was dependent on the underlying distribution of the data being analyzed:

1. One tailed Mann-Whitney U test for comparison of statistical differences between the means between two non-normally distributed and non-homoscedastic distributions.
2. One tailed Welch*’*s t-test for comparison of means between two non-homoscedastic, but normally distributed distributions
3. Brown-Forsythe test for homoscedasticity
4. Shapiro Wilk and D*’*Agostino-Pearson tests for normality.
5. Contingency table chi-squared tests for comparison of frequencies between groups
6. Box and whisker plots
7. Kruskal-Wallis H test for omnibus comparison of means across non-homoscedastic and non-normally distributed groups
8. Pairwise two tailed Mann-Whitney U tests with p-value correction via the Bonferroni-Holm method for post-hoc analysis following a Kruskal-Wallis H test

Statistical significance was evaluated for the 95% confidence level.

## Results

### Analysis of 10-year CVD risk scores allocated by CVD-AI to those who did and did not experience a CV event

In both the UK Biobank testing dataset and the external validation dataset (AREDS 1), the 10-year CV risk scores generated by CVD-AI were significantly higher for patients who had suffered an actual CVD event when compared to patients who did not experience a CVD event. In the Internal validation UK Biobank dataset, the median 10-year CVD risk for those individuals who experienced a CVD was higher than those who did not (4.9% [ICR 2.9-8%] v 2.3% [IQR 4.3-1.3%] P<0.01 one tailed Mann-Whitney U test) Likewise, the mean 10-year CVD risk score for individuals who experienced a CVD event was significantly higher than those who did not (5.8% v 3.3% P<0.01 Welch*’*s t-test). (Figure 2)

**Figure 2:**
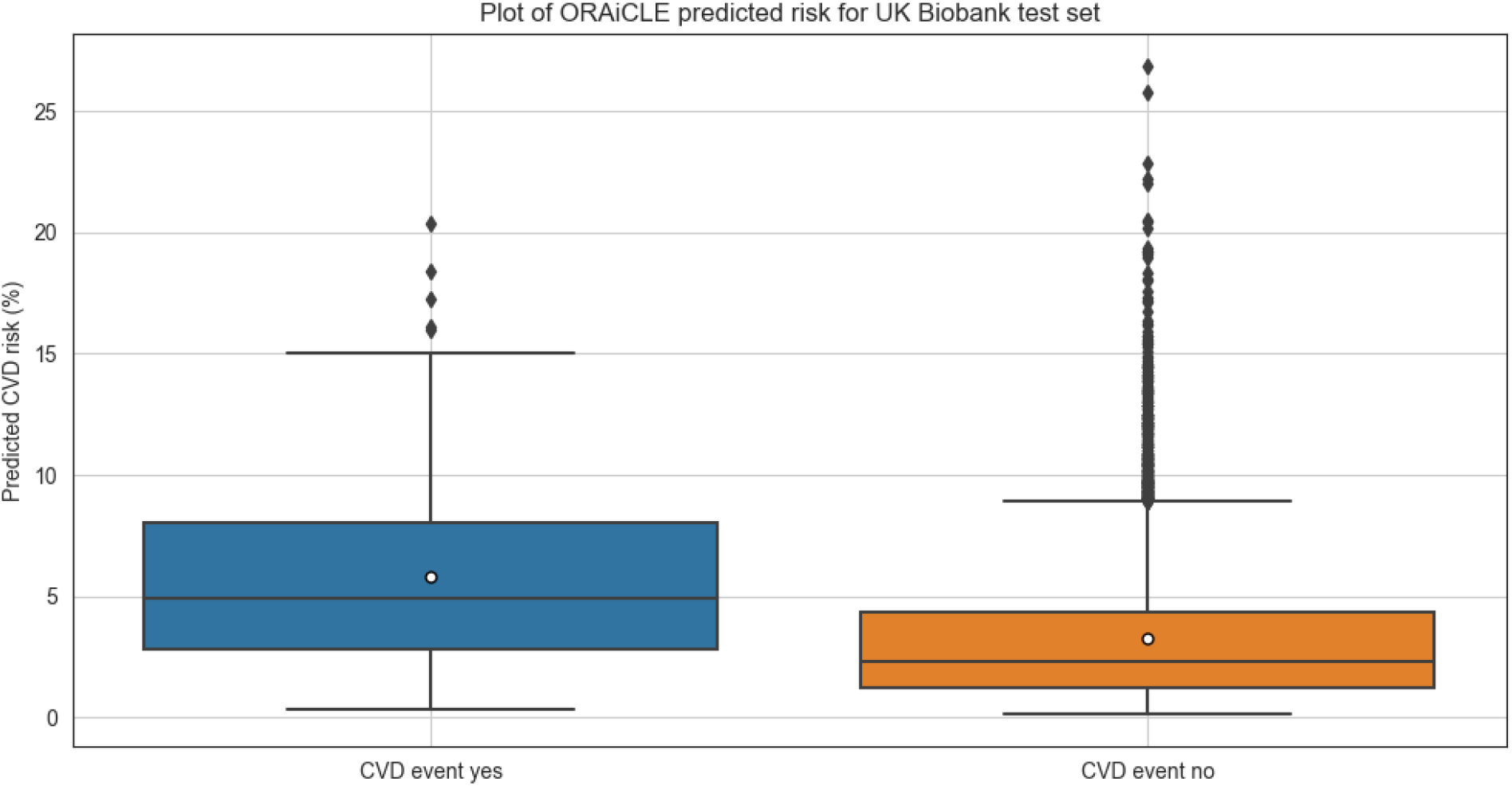
Estimated CVD risk by CVD-AI for people who did vs did not suffer from a CVD event, using the UK Biobank dataset. The center line denotes the median value (50th percentile), while the box contains the 25th to 75th percentiles of dataset. The black whiskers mark the 5th and 95th percentiles, and values beyond these upper and lower bounds are considered outliers. The white circle denotes the mean of the estimated 10-year CVD risk for each category.

Similar results were observed in the AREDS 1 external validation dataset [Figure 3]. The median 10-year CVD risk for those individuals who experienced a CVD event was higher than those who did not (6.2% [ICR 3.2%-12.9%] v 2.2% [IQR 3.9-1.3%] P<0.01 one tailed Mann-Whitney U test) Likewise, the mean 10-year CVD risk score for individuals who experienced a CVD event was 9.0%, v 2.9% for those who did not. (P<0.01 Welch*’*s t-test).

**Figure 3:**
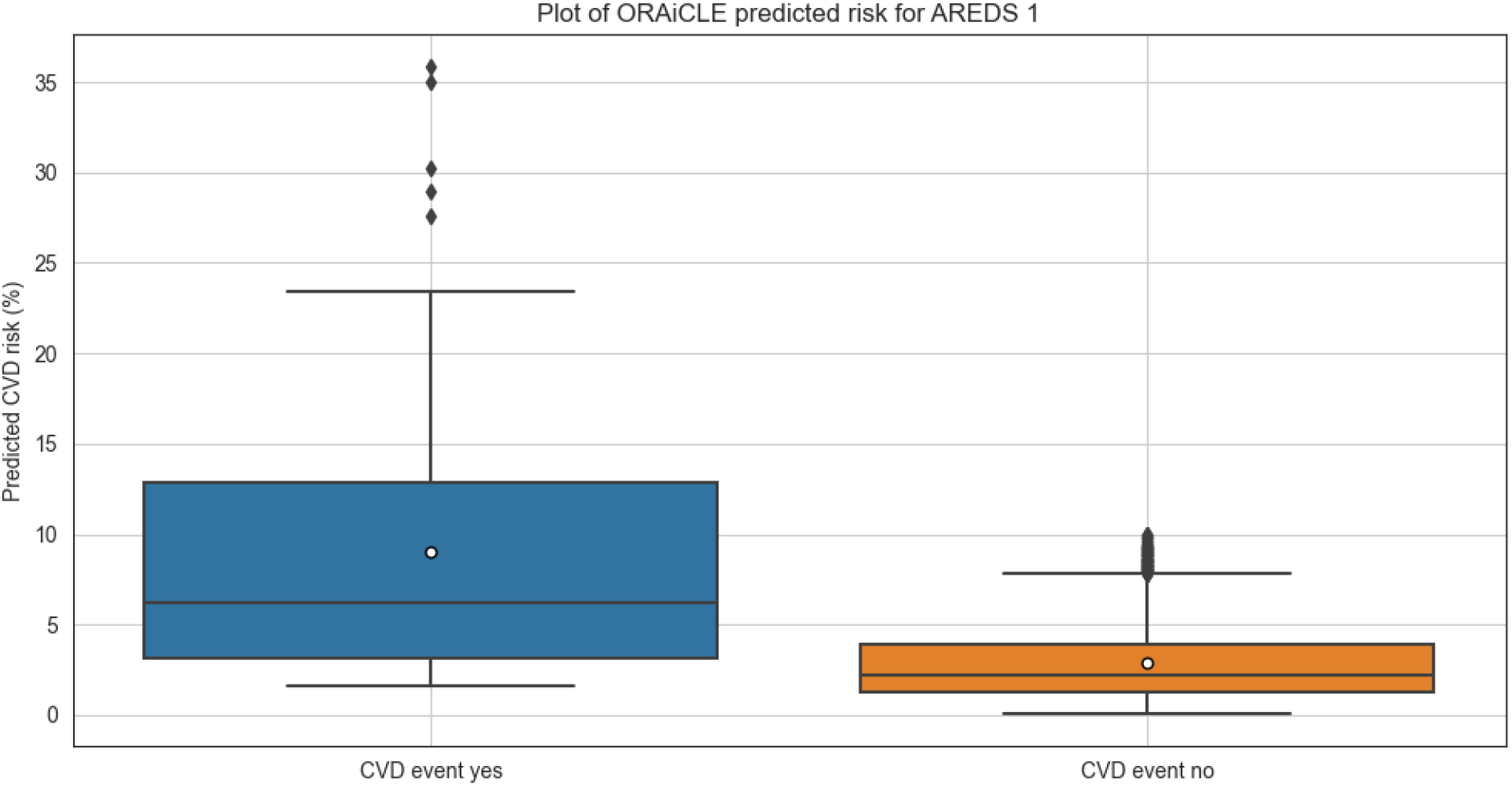
Estimated CVD risk by CVD-AI for people who did vs did not suffer from a CVD event, using the AREDS 1 dataset. The center line denotes the median value (50th percentile), while the box contains the 25th to 75th percentiles of dataset. The black whiskers mark the 5th and 95th percentiles, and values beyond these upper and lower bounds are considered outliers. The white circle denotes the mean of the estimated 10-year CVD risk for each category.

To further evaluate the relevance of the risk scored calculated by the CVD-AI model, arbitrary cutoff point of 5% was chosen based on the box and whisker plots for both the AREDS 1 and the UK Biobank dataset [33-35]. The 5% threshold divided the patients between a low risk (<5%) and elevated risk (>5%) group. 2×2 Chi-squared tests were then carried out based on the two groups. For the UK Biobank testing dataset, the chi-squared tests (X^2^(7,790, 1) = 127.6, p < 0.01) showed that patients who were assigned elevated risk by the CVD-AI algorithm were significantly more likely to have actual CVD events, and patients who were assigned low risk were less likely to have a CVD event. Similar conclusions were reached on the AREDS 1 dataset, with the 2×2 chi-squared test (X^2^(3,162, 1) = 89.1, p < 0.01) showing similar results.

### Metadata analysis

The UK Biobank and AREDS 1 datasets were then recategorized into three groups based on the 10-year CV risk score allocated by CVD-AI. The following thresholds were used: low risk (< 5%), medium risk (5% - 10%) and high risk (> 10%). The numbers of individuals in each category (Low, Medium, High risk), is summarized in table 1.

**Table 1:**
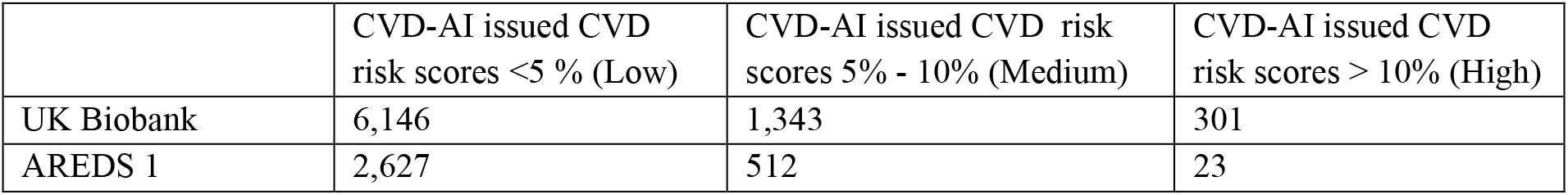
Breakdown of the UK Biobank and AREDS 1 datasets according to the risk score issued by CVD-AI categorized by CVD event risk thresholds: Low, Medium, and High risk.

|  | CVD-AI issued CVD risk scores <5 % (Low) | CVD-AI issued CVD risk scores 5% - 10% (Medium) | CVD-AI issued CVD risk scores > 10% (High) |
| --- | --- | --- | --- |
| UK Biobank | 6,146 | 1,343 | 301 |
| AREDS 1 | 2,627 | 512 | 23 |

Using the UK Biobank test dataset, a metadata analysis was then conducted on factors such as age, HbA1c, systolic blood pressure (taken as the averaged blood pressure between 2 readings), diastolic blood pressure (taken as the averaged blood pressure between 2 readings), BMI, and the total cholesterol to HDL cholesterol (TCHDL) ratio, categorized by the 10-year CV risk score issued by CVD-AI to individuals within these 3 cohorts. Table 2:

**Table 2:** Metadata analysis of the UK Biobank data, categorized by the 10-yr CV risk score allocated by CVD-AI: Low, Medium, and High risk.

|  | Low risk (<5%) |  | Medium risk (5-10%) |  | High risk (>10%) |  | Statistical significance (Yes) |
| --- | --- | --- | --- | --- | --- | --- | --- |
|  | Mean | Standard deviation | Mean | Standard deviation | Mean | Standard deviation |  |
| Age (years) | 55 | 8 | 63 | 5 | 67 | 3 | L – H<br>L – M<br>M – H |
| HbA1C (mmol/mol) | 35 | 6 | 37 | 7 | 38 | 8 | L – H<br>L – M |
| Systolic blood pressure (mmHg) | 135 | 18 | 144 | 18 | 143 | 18 | L – H<br>L – M |
| Diastolic blood pressure (mmHg) | 81 | 9.9 | 84 | 9.7 | 81 | 9.5 | L – H<br>L – M |
| BMI | 27. | 4.9 | 28.0 | 4.4 | 28.0 | 4.1 | L – H<br>L – M |
| TCHDL ratio (No units) | 4.0 | 1.0 | 4.1 | 1.0 | 4.0 | 1.2 | L – H<br>L – M |

|  | Low risk | Medium Risk | High risk | Statistical significance (Yes) |
| --- | --- | --- | --- | --- |
| Proportion of men (%) | 33 | 88 | 98 | L – H<br>L – M<br>M – H |
| Proportion of smokers (%) | 41 | 50 | 59 | L – H<br>L – M<br>M – H |
| Proportion of people with diabetes (%) | 3 | 9 | 13 | L – H<br>L – M<br>M – H |

A similar analysis was conducted on the AREDS 1 dataset for the corresponding fields (where available), again categorized by the 10-year risk score allocated by CVD-AI. Table 3.

**Table 3:** Metadata analysis of the AREDS 1 dataset, categorized by the 10-yr CV risk score allocated by CVD-AI: Low, Medium, and High risk.

|  | Low risk (<5%) |  | Medium risk (5-10%) |  | High risk (>10%) |  | Statistical significance |
| --- | --- | --- | --- | --- | --- | --- | --- |
|  | Mean | Standard deviation | Mean | Standard deviation | Mean | Standard deviation |  |
| Age (years) | 69 | 5 | 70 | 5 | 74 | 5 | L – H<br>L – M<br>M – H |
| Systolic blood pressure (mmHg) | 135 | 17 | 138 | 17 | 144 | 25 | L – M |
| Diastolic blood pressure (mmHg) | 78 | 9 | 78 | 9 | 81 | 10 |  |
| BMI | 27 | 5 | 27 | 5 | 30 | 6 |  |

|  | Low risk | Medium Risk | High risk | Statistical Significance |
| --- | --- | --- | --- | --- |
| Proportion of men (%) | 40 | 56 | 74 | L – H<br>L – M<br>M – H |
| Proportion of smokers (%) | 51 | 59 | 83 | L – H<br>L – M<br>M – H |
| Proportion of people with diabetes (%) | 8 | 9 | 26 | L – H<br>L – M<br>M – H |

### Evaluation of model explainability

To qualitatively evaluate CVD-AI*’*s performance and investigate the *‘*relative contribution*’* of both non-modifiable factors (age, ethnicity, sex) and modifiable factors (HbA1c, blood pressure, smoking and total cholesterol/HDL cholesterol ratio) on the total estimated 10-year risk score. Eight individual case studies from the high-risk groups were created (5 from the UK Biobank and 3 from AREDS 1). The results of the cases and the relative impact of the component risk factors that comprise this overall risk are summarized in Table 4.

**Table 4:** select scenarios from the UK Biobank and AREDS 1 datasets, detailing the patient biometrics, calculated CVD risk and the “relative” contribution of the different components to this risk

|  | UK<br>Biobank 1 | UK<br>Biobank 2 | UK<br>Biobank 3 | UK<br>Biobank 4 | UK<br>Biobank 5 | AREDS<br>1 | AREDS<br>2 | AREDS<br>3 |
| --- | --- | --- | --- | --- | --- | --- | --- | --- |
| Age | 42 | 63 | 62 | 42 | 75 | 65 | 65 | 67 |
| Gender | M | M | M | M | M | F | M | F |
| BMI | 32 | 37 | 39 | 29 | 31 | 28 | 30 | 27 |
| Smoking status | N | Y | Y | N | N | Y | Y | Y |
| Diabetes status | N | N | Y | Y | Y | N | Y | N |
| Systolic blood pressure (mmHg) | 160 | 160 | 161 | 124 | 134 | 142 | 152 | 220 |
| Diastolic blood pressure (mm/Hg) | 105 | 97 | 76 | 84 | 70 | 80 | 90 | 88 |
| HbA1C (mmol/mol) | 37 | 46 | 62 | 55 | 44 | - | - | - |
| Total cholesterol to HDL cholesterol<br>ratio | 7.1 | 4.8 | 3.0 | 5.2 | 2.7 | - | - | - |
| Total Absolute Risk | 5% | 10% | 22% | 24% | 16% | 8% | 13% | 17% |
| Relative Risk Contribution - Age | <10% | 13% | <10% | 60% | 55% | 26% | <10% | 20% |
| Relative Risk Contribution - Gender | 20% | 27% | <10% | 13% | 33% | 13% | <10% | <10% |
| Relative Risk Contribution -<br>Smoking | <10% | <10% | <10% | <10% | <10% | <10% | <10% | 32% |
| Relative Risk Contribution –<br>Systolic BP | 40 % | 17% | <10% | <10% | <10% | 39% | 37% | <10% |
| Relative Risk Contribution -<br>Diabetes | <10% | 12% | 61% | 10% | 10% | <10% | 15% | <10% |
| Relative Risk Contribution -<br>Cholesterol | 20% | 10% | <10% | <10% | <10% | <10% | 40% | 41% |

Table 4: Summary of the demographic and biomarkers for 8 individuals in the Biobank and AREDS 1 datasets and the Absolute and Relative risk scores CVD-AI issued for them.

## Discussion

Previously we have demonstrated that it is possible to train an artificial intelligence (AI) deep learning (DL) algorithm on retinal images to grade diabetic retinopathy and maculopathy for diagnostic, screening and risk assessment purposes [30, 36-45]. In this study we used 110,272 fundus images from a database of 55,118 patients from the UK Biobank and AREDS 1 datasets to train and subsequently test a novel AI platform (CVD-AI) to calculate a 10-year CVD risk score for these individuals. The predicted risk produced by CVD-AI was compared to the actual cardiovascular event rate to determine the relative accuracy of the prediction so obtained. We found that CVD-AI could reliably identify patients at high risk of cardiovascular event, most of whom experienced at least one event according to the UK Biobank or AREDS 1 records.

Our results are in line with other reports which have demonstrated that DL algorithms can use retinal images to predict modifiable CVD risk factors, including diabetes, hypertension, and cholesterol [25, 27, 29, 46-48] and non-modifiable risk factors such as chronological age and gender [24]. However, like the Framingham equations, the algorithms published to date are unable to examine the relative contribution of each of the individual factors that comprise risk as they utilize a statistical method which imposes linearity between the individual parameters used during analysis. Consequently, these models are trained against a single label like cardiovascular event or chronological age. As such they are incapable of identifying the most significant contributors to CVD risk in any given individual as the math underpinning the algorithm do not account for interactions between the variables that comprise the individuals overall risk.

Traditionally, most existing algorithms simply measure success in terms of detection accuracy, where the CVD risk is calculated by conventional equations. For instance, [27, 28, 49, 50] report the outcomes of their algorithms in terms of AUC, regarding successful models as those that have an AUC > 0.70. Although this approach has its merits merely knowing that a model can predict CVD risk with an AUC > 0.70 is of limited value because simply achieving a high level of accuracy does not necessarily mean that the algorithm has learnt what was expected. This is particularly important in the case of biometric data much of which is normally distributed. In data which is normally distributed an algorithm which has simply learnt to assign outputs which are clustered tightly around the mean will, at the population level, be highly accurate. However, in the Real World when presented with individuals whose values fall outside the mean, it will fail to perform. This effect has been elegantly demonstrated by Zhang et al who reported that their algorithm achieved AUC curves of 0.929 for predicting an HbA1C from retinal photographs [51]. However, if the algorithm was actually able to read HbA1C, the accompanying scatter plot of actual v predicted HBA1C would be clustered along the diagonal. As it transpired the predicted values were clustered tightly around the mean HbA1C of the population; along the horizontal meridian. To be sure that an algorithm is performing then the outputs therefore need to be both biologically plausible and clinically meaningful.

To assess the biological and Clinical plausibility of CVD-AI we first evaluated the demographic and biometric data of those individuals allocated to three broad risk categories; low risk (<5%), medium risk (5-10%) and high risk (>10%). These data demonstrated that in both the UK Biobank and AREDS 1 datasets, the demographic and biometric data of the three groups categorized by the results allocated by CVD-AI were largely consistent with traditional cardiovascular risk factors; age gender, smoking, systolic blood pressure and the presence or otherwise of diabetes [8, 34]. However, in addition to the expected traditional metrics other intriguing trends were evident; namely that irrespective of whether the individual had diabetes, HbA1C was significantly and incrementally higher across the 3 groups.

These same trends were observed in the individual case studies. Analysis of these examples again reveal that rising age and male gender were, in the absence of diabetes, the most powerful predictors of cardiovascular risk (Biobank cases 2 & 5, AREDS cases 2 & 3). In contrast, in an older female, systolic hypertension registered more highly than age and gender (AREDS cases 1 & 3). In a younger male patient (Biobank case 1), CVD-AI indicated that the overall risk was low and that in the absence of other CV risk factors, systolic hypertension was the principal factor underpinning this risk. When the individual had diabetes, (Biobank cases 3 & 4, AREDS case 2) blood sugar was one of the principal factors underpinning the individuals CV risk score. However, HbA1C also registered as one of the principle factors underpinning the CV risk score in individuals who did not have diabetes (Biobank case 2 & 5), but whose blood sugar was in the prediabetic range [52]. The finding that HbA1C registers as an important risk factor in patients who don*’*t have diabetes, but whose blood sugar is in the pre diabetic *“*normal*”* range is intriguing. Mean HbA1C was also significantly higher in patients who CVD-AI allocated medium and high-risk scores compared to those allocated low-risk scores. It is well recognized that individuals with prediabetes are at increased risk of not only developing type 2 diabetes, but are also at an increased risk of experiencing a CV event [53, 54]. It is thus possible that CVD-AI is detecting a change within the retina that allows it to discern this subtle signal. As CVD-AI was not trained to predict the HbA1C, this information must instead be derived from as yet unknown changes in the retinal that results from raised; but *“*normal*”* glucose levels. It has recently been reported in prediabetic rat models that elevated, but non diabetic, glucose levels are associated with activation of the TRVP-2 pathway and retinal arteriolar dilation [55]. Although further work in this area is required it is tempting to speculate that a similar process may be at work in the retina of humans with prediabetes.

In recent years newer CVD prediction equations have been developed, based on data gathered from samples that represent the demographics of broader populations in terms of ethnicity, socioeconomic status, and other variables. These developments should lead to an improved predictive power of existing CV risk equations [56]. However, these new CVD risk equations, require the individual being reviewed by a health care profession and rely on data derived from laboratory testing. As such they may be difficult to implement in Health economies where individuals cannot easily access primary care, or these facilities are already over stretched [57]. We have demonstrated that using nothing more than a retinal image, CVD-AI, can evaluate an individual*’*s 10-year cardiovascular risk. Furthermore, and uniquely we believe that we have demonstrated that it possible to train a DL algorithm that is not only able to assess CVD risk at the individual level, but is also able to establish the relative contribution of each risk factor to the overall CVD risk score based on an individual*’*s personal circumstances. If these results can be replicated DL algorithms like CVD-AI offer the potential to significantly improve access to CVD risk prevention strategies. As retinal photographs are routinely captured in Optometric practices it means that they can be deployed without significant additional investment in primary care, a feature which makes these technologies particularly relevant to low-income settings. Finally, AI-based prediction tools that assess risk at the individual level would inform treatment decisions based on the specific needs of an individual, thereby increasing the likelihood of positive health outcomes.

## Conclusion

It is well recognized that many cardiovascular events can be prevented by making treatment recommendations based on an individual*’*s CVD risk profile. However, presently methods used to quantify an individual*’*s risk have only a moderate predictive power and require input from a medical professional augmented with laboratory tests. In this paper, we demonstrate that our DL algorithm CVD-AI, using a retinal image as the sole input, is capable of assessing both the 10-year risk of an individual experiencing a cardiovascular event and identify the relative components from which this risk score is derived.

## Data Availability

All data produced are available online, through the original data guardians, and applications are received from the public

https://www.ncbi.nlm.nih.gov/projects/gap/cgi-bin/study.cgi?study_id=phs000001.v3.p1

https://www.ukbiobank.ac.uk/enable-your-research/apply-for-access#:~:text=If%20you%20have%20already%20registered,Biobank%20data%2Dfields%20you%20require

## Supplementary Tables

**Supplementary Table 1:** the makeup of the UK BioBank dataset used here

|  | Mean | Std |
| --- | --- | --- |
| Age | 56.8 | 8.26 |
| Systolic Blood Pressure (mmHg) | 136.96 | 18.4 |
| Diastolic Blood Pressure (mmHg) | 81.69 | 9.93 |
| Hba1c (mmol/mol) | 35.88 | 6.29 |
| BMI | 27.2 | 4.7 |
|  | Male | Female |
| Gender | 23,482 | 28,473 |
|  | TRUE | FALSE |
| Smoking | 22,366 | 29,589 |
| CVD event | 264 | 7526 |

**Supplementary Table 2:**
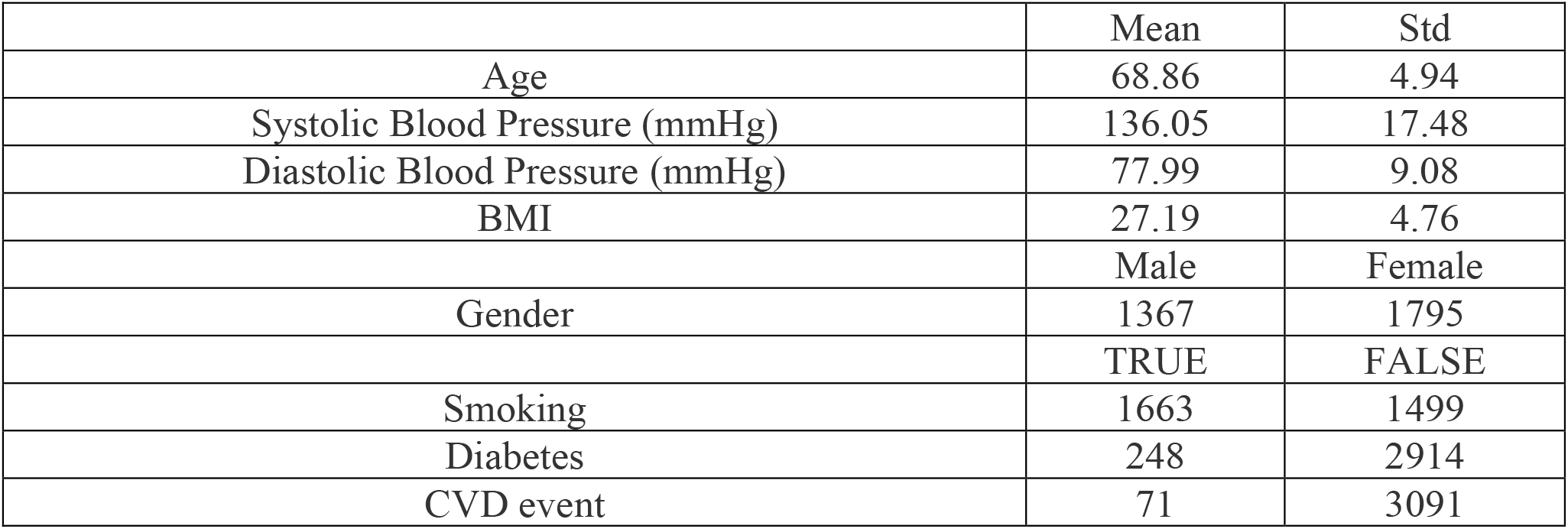
the makeup of the AREDS 1 dataset used here

## References

1. Control, C.f.D. and Prevention, Underlying cause of death, 1999–2018. CDC WONDER Online Database. Atlanta, GA: Centers for Disease Control and Prevention, 2018.

2. Almourani, R., et al., Diabetes and cardiovascular disease: an update. Current diabetes reports, 2019. 19(12): p. 1–13.

3. Kjeldsen, S.E., Hypertension and cardiovascular risk: General aspects. Pharmacological research, 2018. 129: p. 95–99.

4. Alloubani, A., R. Nimer, and R. Samara, Relationship between hyperlipidemia, cardiovascular disease and stroke: a systematic review. Current Cardiology Reviews, 2021. 17(6): p. 52–66.

5. Kondo, T., et al., Effects of tobacco smoking on cardiovascular disease. Circulation Journal, 2019. 83(10): p. 1980–1985.

6. Lloyd-Jones, D.M., et al., Use of risk assessment tools to guide decision-making in the primary prevention of atherosclerotic cardiovascular disease: a special report from the American Heart Association and American College of Cardiology. Circulation, 2019. 139(25): p. e1162–e1177.

7. Mahmood, S.S., et al., The Framingham Heart Study and the epidemiology of cardiovascular disease: a historical perspective. The lancet, 2014. 383(9921): p. 999–1008.

8. Kavousi, M., et al., Comparison of application of the ACC/AHA guidelines, Adult Treatment Panel III guidelines, and European Society of Cardiology guidelines for cardiovascular disease prevention in a European cohort. Jama, 2014. 311(14): p. 1416–1423.

9. DeFilippis, A.P., et al., An analysis of calibration and discrimination among multiple cardiovascular risk scores in a modern multiethnic cohort. Annals of internal medicine, 2015. 162(4): p. 266–275.

10. Comín, E., et al., Estimating cardiovascular risk in Spain using different algorithms. Revista Española de Cardiología (English Edition), 2007. 60(7): p. 693–702.

11. Goff, D., D. Lloyd-Jones, and G. Bennett, 2013 ACC/AHA Guideline on the Assessment of Cardiovascular Risk: A Report of the American College of Cardiology/American Heart Association Task Force on Practice Guidelines. J Am Coll Cardiol 2013 Nov 12 [E-pub ahead of print. Journal of the American College of Cardiology, 2014. 63(25).

12. D’Agostino, R.B., et al., Validation of the Framingham coronary heart disease prediction scores: results of a multiple ethnic groups investigation. Jama, 2001. 286(2): p. 180–187.

13. Beswick, A., et al., A systematic review of risk scoring methods and clinical decision aids used in the primary prevention of coronary heart disease (supplement). 2011.

14. Brindle, P., et al., Accuracy and impact of risk assessment in the primary prevention of cardiovascular disease: a systematic review. Heart, 2006. 92(12): p. 1752–1759.

15. Eichler, K., et al., Prediction of first coronary events with the Framingham score: a systematic review. American heart journal, 2007. 153(5): p. 722-731. e8.

16. Damen, J.A., et al., Performance of the Framingham risk models and pooled cohort equations for predicting 10-year risk of cardiovascular disease: a systematic review and meta-analysis. BMC medicine, 2019. 17(1): p. 1–16.

17. McEwan, P., et al., Evaluating the performance of the Framingham risk equations in a population with diabetes. Diabetic medicine, 2004. 21(4): p. 318–323.

18. Francula-Zaninovic, S. and I.A. Nola, Management of measurable variable cardiovascular disease’risk factors. Current cardiology reviews, 2018. 14(3): p. 153–163.

19. Schisterman, E.F. and B.W. Whitcomb, Coronary age as a risk factor in the modified Framingham risk score. BMC medical imaging, 2004. 4(1): p. 1–9.

20. Hira, R.S., et al., Frequency and practice-level variation in inappropriate aspirin use for the primary prevention of cardiovascular disease: insights from the National Cardiovascular Disease Registry’s Practice Innovation and Clinical Excellence registry. Journal of the American College of Cardiology, 2015. 65(2): p. 111–121.

21. Mochari, H., et al., Cardiovascular disease knowledge, medication adherence, and barriers to preventive action in a minority population. Preventive cardiology, 2007. 10(4): p. 190–195.

22. Armstrong, G.W. and A.C. Lorch, A (eye): a review of current applications of artificial intelligence and machine learning in ophthalmology. International ophthalmology clinics, 2020. 60(1): p. 57–71.

23. Miyazawa, A.A., Artificial intelligence: the future for cardiology. Heart, 2019. 105(15): p. 1214–1214.

24. Zhu, Z., et al., Retinal age gap as a predictive biomarker for mortality risk. British Journal of Ophthalmology, 2022.

25. Cheung, C.Y., et al., A deep-learning system for the assessment of cardiovascular disease risk via the measurement of retinal-vessel calibre. Nature biomedical engineering, 2021. 5(6): p. 498–508.

26. Chang, J., et al., Association of cardiovascular mortality and deep learning-funduscopic atherosclerosis score derived from retinal fundus images. American Journal of Ophthalmology, 2020. 217: p. 121–130.

27. Poplin, R., et al., Prediction of cardiovascular risk factors from retinal fundus photographs via deep learning. Nature Biomedical Engineering, 2018. 2(3): p. 158.

28. Ma, Y., et al., Development and validation of a deep learning algorithm using fundus photographs to predict 10-year risk of ischemic cardiovascular diseases among Chinese population. medRxiv, 2021.

29. Velasco, A.V., et al., Decreased retinal vascular complexity is an early biomarker of MI supported by a shared genetic control. medRxiv, 2021.

30. Xie, L., et al., Towards implementation of AI in New Zealand national diabetic screening program: Cloud-based, robust, and bespoke. Plos one, 2020. 15(4): p. e0225015.

31. Anderson, K.M., et al., Cardiovascular disease risk profiles. American heart journal, 1991. 121(1): p. 293–298.

32. Nielsen, I.E., et al., Robust explainability: A tutorial on gradient-based attribution methods for deep neural networks. IEEE Signal Processing Magazine, 2022. 39(4): p. 73–84.

33. Goff Jr, D.C., et al., 2013 ACC/AHA guideline on the assessment of cardiovascular risk: a report of the American College of Cardiology/American Heart Association Task Force on Practice Guidelines. Circulation, 2014. 129(25_suppl_2): p. S49–S73.

34. Stone, N.J., et al., 2013 ACC/AHA guideline on the treatment of blood cholesterol to reduce atherosclerotic cardiovascular risk in adults: a report of the American College of Cardiology/American Heart Association Task Force on Practice Guidelines. Journal of the American College of Cardiology, 2014. 63(25 Part B): p. 2889–2934.

35. Bibbins-Domingo, K., et al., Statin use for the primary prevention of cardiovascular disease in adults: US Preventive Services Task Force recommendation statement. Jama, 2016. 316(19): p. 1997–2007.

36. Aaron Yap, L.H., Eileen Chen, Ehsan Vaghefi, David Squirrell, Patients’ perceptions of artificial intelligence in diabetic eye screening, in 2022 RANZCO. 2022, The Royal Australian and New Zealand College of Ophthalmologists: Brisbane - Australia.

37. Chu, A., et al., Essentials of a robust deep learning system for diabetic retinopathy screening: a systematic literature review. Journal of Ophthalmology, 2020. 2020.

38. Hill, S., et al., Risk Factors for Progression to Referable Diabetic Eye Disease in People With Diabetes Mellitus in Auckland, New Zealand: A 12-Year Retrospective Cohort Analysis. The Asia-Pacific Journal of Ophthalmology, 2021. 10(6): p. 579–589.

39. Li Xie, E.V., Song Yang, David Han, John Marshall, David Squirrell, Automation of macular degeneration classification in the AREDS dataset, using a novel neural network design - (in press). Ophthalmology Science, 2022. 21(00226).

40. Nishanthan Ramachandran, O.S., Ehsan Vaghefi, Sophie Hill, Graham A Wilson, David Squirrell, Evaluation of the prevalence of non-diabetic eye disease detected at first screen from a single region diabetic retinopathy screening program; a cross-sectional cohort study in Auckland, New Zealand. BMJ Open, 2021. In-Press.

41. Vaghefi, E., et al., Multimodal retinal image analysis via deep learning for the diagnosis of intermediate dry age-related macular degeneration: a feasibility study. Journal of ophthalmology, 2020. 2020.

42. Vaghefi, E., et al., Detection of smoking status from retinal images; a Convolutional Neural Network study. Scientific reports, 2019. 9(1): p. 1–9.

43. Vaghefi, E., et al., A multi-center prospective evaluation of THEIA to detect diabetic retinopathy (DR) and diabetic macular edema (DME) in the New Zealand screening program. arXiv preprint 2106.12979, 2021.

44. Vaghefi, E., et al., THEIA™ development, and testing of artificial intelligence-based primary triage of diabetic retinopathy screening images in New Zealand. Diabetic Medicine, 2021. 38(4): p. e14386.

45. Vaghefi, E., et al., A multi-centre prospective evaluation of THEIA™ to detect diabetic retinopathy (DR) and diabetic macular oedema (DMO) in the New Zealand screening program. Eye, 2022: p. 1–7.

46. Wagner, S.K., et al., Insights into systemic disease through retinal imaging-based oculomics. Translational Vision Science & Technology, 2020. 9(2): p. 6–6.

47. Tapp, R.J., et al., Associations of retinal microvascular diameters and tortuosity with blood pressure and arterial stiffness: United Kingdom Biobank. Hypertension, 2019. 74(6): p. 1383–1390.

48. Chandra, A., et al., The association of retinal vessel calibres with heart failure and long-term alterations in cardiac structure and function: the Atherosclerosis Risk in Communities (ARIC) Study. European Journal of Heart Failure, 2019. 21(10): p. 1207–1215.

49. Huang, F., et al., Predicting CT-Based Coronary Artery Disease Using Vascular Biomarkers Derived from Fundus Photographs with a Graph Convolutional Neural Network. Diagnostics, 2022. 12(6): p. 1390.

50. Zhang, L., et al., Prediction of hypertension, hyperglycemia and dyslipidemia from retinal fundus photographs via deep learning: A cross-sectional study of chronic diseases in central China. PloS one, 2020. 15(5): p. e0233166.

51. Zhang, K., et al., Deep-learning models for the detection and incidence prediction of chronic kidney disease and type 2 diabetes from retinal fundus images. Nature Biomedical Engineering, 2021. 5(6): p. 533–545.

52. Yudkin, J.S., “Prediabetes”: Are There Problems With This Label? Yes, the Label Creates Further Problems! Diabetes Care, 2016. 39(8): p. 1468–1471.

53. Cai, X., et al., Association between prediabetes and risk of all cause mortality and cardiovascular disease: updated meta-analysis. Bmj, 2020. 370.

54. Beulens, J., et al., Risk and management of pre-diabetes. European journal of preventive cardiology, 2019. 26(2_suppl): p. 47–54.

55. O’Hare, M., et al., Loss of TRPV2-mediated blood flow autoregulation recapitulates diabetic retinopathy in rats. JCI insight, 2022. 7(18).

56. Lira, M., et al., Validity of an adaptation of the framingham cardiovascular risk function: comparison of predicted and observed outcomes with international risk models. 2018.

57. Bitton, A. and T. Gaziano, The Framingham Heart Study’s impact on global risk assessment. Progress in cardiovascular diseases, 2010. 53(1): p. 68–78.

